# Part 1: Examining heterogeneity of treatment effects in transcranial direct current stimulation for knee osteoarthritis pain and symptoms

**DOI:** 10.1101/2025.06.09.25329205

**Authors:** Chiyoung Lee, Xiaoxiao Sun, Juyoung Park, Chen X. Chen, Christine Pellegrini, Nan-kuei Chen, David O. Garcia, Heewon Kim, C. Kent Kwoh, Hyochol Ahn

## Abstract

**Background:** Although heterogeneity of treatment effects (HTE) is commonly observed in clinical trials, it has received little attention in studies on transcranial direct current stimulation (tDCS). This study aimed to identify the presence of HTE in tDCS treatment among participants with symptomatic knee osteoarthritis (KOA) and to explore participant characteristics associated with this heterogeneity.

**Methods:** This exploratory secondary analysis of a randomized clinical trial included 120 participants with symptomatic KOA who received 15 daily sessions of home-based 2-mA active or sham tDCS (20 minutes per session) over three weeks. First, we used a multi-trajectory latent class growth analysis to identify distinct subgroups based on the longitudinal trajectories of KOA pain and symptoms from baseline to three months postintervention, capturing differential responses to tDCS. We then performed bivariate analyses to examine associations between trajectory groups and baseline demographic, clinical, and quantitative sensory testing characteristics.

**Results:** In the active tDCS group, two distinct trajectories emerged: “low initial symptoms with significant improvement” (high responders; *n* = 28) and “high initial symptoms with minimal improvement” (low responders; *n* = 32). Compared to high responders, low responders had a higher body mass index (*p* = .040), lower educational attainment (*p* = .013), and greater pain catastrophizing *(p* < .000). Low responders also exhibited lower pressure pain thresholds at both the medial knee (*p* = .009) and trapezius (*p* = .002), higher punctate mechanical pain at both the patella (*p* = .013) and hand (*p* = .016), lower conditioned pain modulation at 30 seconds (*p* = .008) and 60 seconds (*p* < .000), and higher cold pain intensity (*p* = .003) at baseline. No notable HTE was observed in the sham tDCS group.

**Conclusion:** Participants exhibited varying responses to active tDCS. The characteristics associated with HTE may inform the development of personalized stimulation protocols. Further research is needed to investigate potential HTE in the sham tDCS group and refine strategies to address placebo-related effects.

## 1. Introduction

The public health burden of osteoarthritis in the U.S. is expected to rise substantially due to the aging population.^1^ Knee osteoarthritis (KOA) is the most prevalent form of arthritis, affecting more than 14 million older adults in the U.S. and leading to chronic pain and reduced quality of life.^2,3^ Since KOA is currently incurable, its management primarily focuses on pain relief and functional improvement. Although pharmacological treatments for KOA show promise, they often cause side effects, highlighting the need for novel nonpharmacological approaches.

Emerging research suggests that pain-related brain mechanisms are altered in individuals with KOA pain and that altered central pain processing is a key driver of joint pain and dysfunction.^4,5^ This has led to growing interest in nonpharmacological strategies targeting central pain mechanisms, particularly transcranial direct current stimulation (tDCS).^6^ The pain-modulating effects of tDCS are believed to result from a complex interplay of mechanisms, including the modulation of cortical excitability, activation of endogenous pain control systems, induction of neural plasticity, and regulation of neurotransmitters.^6^

The efficacy of tDCS in alleviating KOA pain and symptoms has been demonstrated in several clinical trials, with the primary motor cortex (M1) being the most commonly targeted brain area for stimulation.^7–10^ Conventionally, intervention efficacy is assessed based on the average treatment response. However, differential responses to treatments, often referred to as the “heterogeneity of treatment effects” (HTE), are inherent in intervention outcomes.^11^ Not all patients in treatment groups respond in the same way or to the same extent, with some showing no meaningful improvement—an outcome typically classified as “low responders” (low efficacy) compared to “high responders” (high efficacy). These findings raise concerns, particularly when certain individuals fail to benefit from active treatment, as the implications for clinical decision-making could be substantial. Additionally, some patients in the sham control groups may exhibit a “treatment response” despite receiving no active intervention, underscoring the difficulty in interpreting tDCS outcomes. To date, HTE in active and sham tDCS have rarely been empirically examined in tDCS for KOA.

Several factors may contribute to the HTE of tDCS. Pain perception and response to treatment vary among individuals, making personalized care particularly relevant. According to de Rooij et al.,^12^ HTE in multidisciplinary treatment for chronic widespread pain was associated with gender and baseline levels of pain and anxiety. Similarly, Boonstra et al.^13^ reported that HTE in multidisciplinary pain rehabilitation was linked to baseline levels of pain, disability, and depression in patients with chronic musculoskeletal pain. Given the considerable phenotypic heterogeneity among individuals with KOA, a one-size-fits-all treatment approach is unlikely to be effective. Treatment planning must account for individual variability in contributing factors and symptomatic burden. Such insights may help inform personalized brain stimulation protocols, ensuring that interventions are tailored to each patient’s specific characteristics, ultimately maximizing therapeutic benefits and minimizing ineffective treatments.^14^

This exploratory study aimed to identify the presence of HTE in tDCS treatment among participants with symptomatic KOA and to examine participant characteristics associated with this heterogeneity. First, we classified individuals and assessed HTE based on their longitudinal KOA pain and symptom patterns from baseline to postintervention follow-up; this approach accounted for differences in the time course of symptomatic change to determine differential responses to tDCS. Associations between the identified trajectory groups and baseline demographic, clinical, and quantitative sensory testing (QST) characteristics were then examined. The sham tDCS group was analyzed separately to assess potential HTE, providing insights into subgroup susceptibility to placebo effects and strategies to mitigate them.

## 2. Methods

### 2.1. Design

This secondary analysis utilized data from a double-blind, randomized, sham-controlled phase II pilot clinical trial with a parallel group design. The trial was registered on ClinicalTrials.gov (NCT04016272). It involved a three-week home-based tDCS program, monitored in real time via secure videoconferencing. A total of 120 participants, all of whom provided informed written consent, were randomly assigned to either active or sham tDCS groups, with 60 participants in each group (**Figure 1**). Randomization was conducted in the order of study enrollment using a pre-generated randomization list created with SAS© software, Version 9.4 (SAS Institute Inc., Cary, NC) by a statistician uninvolved in the clinical aspects of the trial. Covariate-adaptive randomization was applied to ensure balance across the groups.

**Figure 1.**
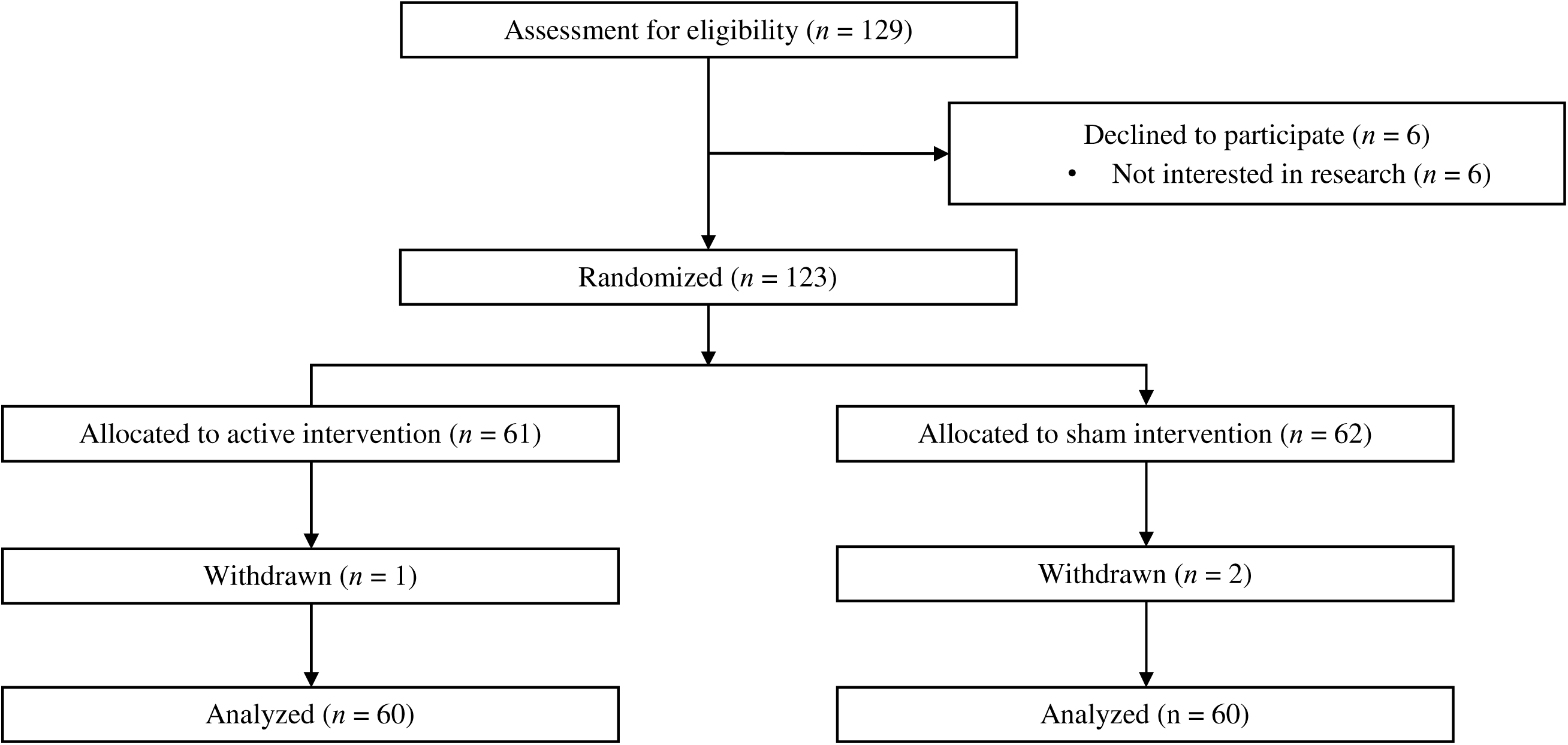
**Participant flow diagram (*n* =120)** *Note.* “Declined” meant that participants came to baseline visit (signed informed consent) but did not start any intervention. For this study, among 6 declined subjects, 4 subjects signed informed consent at the baseline but declined to study participation; 2 subjects did not show up at the baseline.

### 2.2. Participants

Eligible participants were adults aged 50–85 years who met the following criteria: (1) symptomatic KOA diagnosed based on the American College of Rheumatology criteria, with radiographic assessments determining osteoarthritis severity using Kellgren-Lawrence scores, (2) KOA pain experienced over the past three months, with an average pain score of at least 30 on a 0–100 numerical rating scale, (3) the ability to speak and read English, and (4) no plans to modify their pain medication regimen during the study period. According to the American College of Rheumatology criteria for classifying osteoarthritis, participants were required to meet at least three of six criteria: age >50 years, stiffness lasting <30 minutes, presence of crepitus, bony tenderness, bony enlargement, and absence of palpable warmth.

Participants were excluded if they had medical conditions that could affect the interpretation of results, pose safety risks during assessments or tDCS procedures, or interfere with the successful completion of the study protocol. Specifically, individuals were excluded if they: (1) had undergone prosthetic knee replacement or non-arthroscopic surgery on the affected knee; (2) had a history of brain surgery, brain tumor, seizure, stroke, or intracranial metal implants; (3) were diagnosed with systemic rheumatic diseases, such as rheumatoid arthritis, systemic lupus erythematosus, or fibromyalgia; (4) had alcohol or substance use disorders; (5) were receiving treatment with sodium channel blockers, calcium channel blockers, or N-methyl-D-aspartate receptor antagonists receptor antagonists; (6) had cognitive impairments (Mini-Mental Status Exam Score ≤ 23) that could hinder their understanding of study procedures; (7) were pregnant or breastfeeding; (8) had a history of psychiatric hospitalization within the past year; or (9) lacked internet access.

Participants were recruited from Texas through a multi-faceted approach, including advertisements at local healthcare institutions, community flyers, and direct recruitment from a regional orthopedic clinic.

### 2.3. Intervention

**Active tDCS.** The tDCS device used in the study was the Soterix 1×1 tDCS mini-CT Stimulator (Soterix Medical Inc., New York), equipped with headgear and 5 × 7 cm saline-soaked sponge electrodes. The electrodes were securely affixed to the custom headgear, designed to facilitate precise placement and minimize setup errors.^15^ The single-position headgear, with clearly labeled sponge locations, ensured accurate and consistent electrode positioning. The anode was placed over the M1, while the cathode was positioned on the contralateral supraorbital area. Stimulation over the M1 is critical for motor control and central pain processing.^16^ Each session delivered a constant 2-mA (subthreshold intensity) for 20 minutes, with a 30-second ramp-up and ramp-down period to maintain effective blinding.^17^ Participants completed a total of 15 sessions over three weeks, with five sessions per week.

Participants received comprehensive training on the the tDCS device during their baseline visit. Upon confirmation of their understanding by the research staff, they were provided with a tDCS device, an organized daily device kit, and a pictorial manual containing detailed instructions. Stimulation sessions could only be initiated after participants obtained a unique unlock code from the research team. Once proper contact quality was established, the sessions began with device settings locked to prevent any adjustments. To ensure double-blinding, the device required a five-digit single-use activation code, preprogramed for a specific stimulation sequence. Both participants and experimenters remained blinded to group assignments. Upon entering the unlock code, the device displayed a 20-minute countdown timer for the session. At the session’s conclusion, the device automatically powered off, and participants were instructed to remove and discard the sponges before securely storing the equipment for subsequent use

**Sham tDCS.** The sham stimulation setup was identical to that of active tDCS to maintain blinding. The stimulator was only activated for 30 seconds at the beginning and end of each session, stimulating the initial and concluding sensory experience of active tDCS without delivering sustained current. This method effectively preserved blinding by ensuring participants could not distinguish between active and sham stimulation.^18^

### 2.4. Measurements

#### 2.4.1. KOA Pain and Symptoms

KOA pain and symptoms were assessed at baseline, after five intervention sessions (Day 5), after an additional five sessions (Day 10), and after the final set of five sessions (Day 15). Follow-up evaluations occurred at 1 month (Day 45), 2 months (Day 75), and 3 months (Day 105) post-intervention.

**Numeric Rating Scale (NRS).** Participants rated their average knee pain over the past 24 hours using the NRS, a scale ranging from 0 (no pain) to 100 (worst pain imaginable). The NRS is a reliable and well-validated measure that is widely used for its sensitivity in detecting pain-related changes among adults with KOA.^19^

**The Western Ontario and McMaster Universities Osteoarthritis Index (WOMAC).** The WOMAC is a self-administered composite questionnaire designed to assess pain, stiffness, and physical function.^20^ It is the most widely recommended disease-specific instrument for core set assessments in KOA clinical trials, as established by the Osteoarthritis Society Task Force.^21^ The pain subscale consists of five questions evaluating pain severity during walking, stair climbing, sleeping, resting, and standing, rated on a 0–4 Likert scale (0 = no pain, 4 = extreme pain). The stiffness subscale includes two questions—“How severe is your stiffness after first awakening in the morning?” and “How severe is your stiffness after sitting, lying, or resting later in the day?”—both rated on a 0–4 Likert scale (0 = no stiffness, 4 = extreme stiffness). The physical function subscale consists of 17 questions assessing everyday activities, such as stair use, standing up from a seated position, bending, and walking. Each participant’s responses to these functional disability questions are rated on a 5-point scale (0 = no difficulty, 4 = extreme difficulty). The responses for each subscale are aggregated to generate composite scores for each dimension.

#### 2.4.2. Baseline Characteristics

##### 2.4.2.1. Demographic and Clinical Characteristics

Demographic characteristics, including age (continuous), gender (male *vs.* female), body mass index (BMI; kg/m²), race (white *vs*. non-white), education (high school or less *vs.* college or more), and marital status (married/partnered *vs.* non-married/unpartnered), as well as clinical characteristics, such as the index knee (the most affected knee), Kellgren-Lawrence score, and the average duration of osteoarthritis (months), were collected. Pain catastrophizing was assessed using the Pain Catastrophizing Scale (PCS), a 13-item measure designed to evaluate catastrophic thinking related to pain across three dimensions: rumination (four items), magnification (three items), and helplessness (six items).^22,23^ Each item was rated on a five-point scale ranging from “not at all” (0) to “all the time” (4). The total score was calculated by summing the raw scores across all items. The PCS demonstrated adequate internal consistency, with subscale alphas ranging from 0.66 to 0.87 (α for all items = 0.87).^22^ Its sensitivity to psychosocial interventions for chronic pain has been well established.^24^

##### 2.4.2.1 QST Procedures

QST is a research and clinical method used to measure responses to sensory and painful stimulations, providing insight into altered pain sensitivity or modulation.^25,26^ A multimodal QST battery was administered, including heat pain threshold (HPTh), heat pain tolerance (HPTo), pressure pain threshold (PPTh), punctate mechanical pain, temporal summation of pain (TSP), conditioned pain modulation (CPM), and cold pain. TSP, which involves the repeated administration of noxious stimuli at a fixed intensity, induces a gradual increase in pain sensation and is believed to reflect endogenous pain-excitatory mechanisms.^27^ CPM assesses the function of the endogenous pain-inhibitory pathway, commonly referred to as the “pain inhibits pain” mechanism.^28^ To minimize carryover effects, the sequence of heat and mechanical testing was randomized and counterbalanced, while CPM was always administered last.

**Thermal Testing Procedures.** Contact heat stimuli were delivered using a computer-controlled TSA-II NeuroSensory Analyzer (Medoc Ltd., Ramat Yishai, Israel) to assess HPTh and HPTo at both the index knee and the ipsilateral ventral forearm, employing an ascending method of limits. To prevent sensitization or habituation of cutaneous receptors, the thermode position was rotated among three locations between trials at each body site. Starting at a baseline temperature of 32°C, the thermode temperature increased at a rate of 0.5°C per second until participants pressed a button on a handheld device. Participants were instructed to press the button when they first perceived the heat as painful to measure HPTh and when the heat became intolerable to assess HPTo. Three HPTh trials were conducted at the first test site, followed by three HPTo trials. The same sequence was then repeated at the second test site, with a 5-minute rest period between sites. The average of the three trials was calculated for each participant to determine overall HPTh and HPTo temperatures for analysis.

**Mechanical Testing Procedures.** PPTh was evaluated using a handheld digital pressure algometer (Wagner, Greenwich, Connecticut, USA) to apply blunt mechanical pressure to deep tissues, including muscles and joints. The pressure was increased at a constant rate of 0.3 kgf/cm² per second to determine PPTh at two locations: the medial side of the index knee and the trapezius muscle. The order of testing sites was randomized and counterbalanced. Participants were instructed to indicate the point at which the sensation “first became painful” and at which point the applied pressure was recorded. The average of three trials at each site was calculated to determine the PPTh for that location.

After assessing PPTh, participants underwent testing to evaluate cutaneous mechanical sensitivity to punctate stimuli on both the index patella and back of the ipsilateral hand. A calibrated nylon monofilament delivering a target force of 300 g was applied 10 times at a rate of one contact per second, with participants providing verbal pain intensity ratings on a scale from 0 (no pain sensation) to 100 (the most intense pain imaginable). Pain ratings from the two trials were averaged to punctate mechanical pain at each site. For TSP assessment, participants first rated the pain intensity from a single application of the monofilament. They then rated the maximum pain intensity experienced during a series of 10 contacts administered at a rate of one contact per second. Temporal summation of pain was calculated by subtracting the pain rating for the single stimulus from the rating for the series of 10 stimuli at each site.

**CPM.** Ten minutes after assessing thermal or mechanical pain, CPM was evaluated by calculating the change in PPTh during the cold pressor task relative to baseline PPTh. Baseline PPTh measurements were obtained immediately before participants immersed their hands in a cold water bath maintained at 12°C. Thirty seconds after immersion, participants rated their cold pain intensity on a scale from 0 to 100, followed by a second PPTh measurement. Participants were instructed to keep their hands submerged for as long as tolerable for up to a maximum of one minute. Upon hand removal, the final PPTh measurements were recorded. CPM at 30 and 60 seconds was calculated by subtracting baseline PPTh from PPTh at 30 seconds and PPTh at 60 seconds, respectively. The 12°C temperature was selected based on prior studies involving middle-aged and older adults with KOA, as it was found to induce moderate yet tolerable pain in most participants.^29^ The water was continuously circulated and maintained at a stable temperature using a refrigeration unit (Neslab, Portsmouth, New Hampshire, USA). An increase in PPTh following cold water immersion indicated pain inhibition.

### 2.5. Statistical Analysis

This study examined HTE through a subgroup analysis that compared effects among participant groups defined as “one variable at a time” (e.g., male vs. female). The active and sham tDCS groups were analyzed separately to assess potential HTE.

First, multi-trajectory latent class growth analysis^30^ was conducted using the SAS procedure *Proc Traj* to identify distinct latent groups with similar trajectories based on jointly modeled KOA pain and symptom parameters from baseline to the three-month post-intervention follow-up. This method allows for the simultaneous modeling of KOA pain and symptom parameters, maximizing the use of available data, particularly when strong interrelationships or collinearity exists among symptoms (e.g., pain, stiffness, and physical function). *Proc Traj* was selected for its ability to identify developmental trajectories, its modeling flexibility and its capacity to handle missing data. The model incorporates all available observations and assumes that missing data occur at random. Both linear and quadratic trend models were tested, and models with one to three latent groups were analyzed and compared. The optimal number of multi-trajectory groups was determined using several fit criteria, including the Akaike information criterion (AIC), Bayesian information criterion (BIC), homogeneity, latent class separation, interpretability, and group proportion, with the latter two being the most critical factors. Lower AIC and BIC values indicated a better-fitting model. Each group had to represent at least 10.0% of the sample to ensure meaningful interpretation and facilitate subsequent analyses, as smaller groups might lack interpretability or be difficult to replicate.^31^

Second, the relationship between the identified trajectory groups and (a) demographic, (b) clinical, and (c) QST features was evaluated. Categorical variables were analyzed using the chi-square test or Fisher’s exact test, while continuous variables were assessed using the t-test or Mann–Whitney U test, depending on data normality. All analyses were conducted using SAS© software, Version 9.4 (SAS Institute Inc., Cary, North Carolina, USA), with statistical significance set at *p* <.05.

## 3. Results

The active tDCS group had a mean age of 65.32 ± 8.41 years, with 66.7% female participants, while the sham tDCS group had a mean age of 66.60 ± 8.43 years, with 70% female participants. White participants comprised 43.3% of the active tDCS group and 53.3% of the sham tDCS group. Additional details on the baseline characteristics of each treatment group are provided in **Supplemental Table 1**. The best-fitting models for most longitudinal latent classes were characterized by linear slopes (**Supplemental Table 2**). The intervention was well tolerated by participants, with no significant side effects reported. All participants completed the prescribed number of sessions, resulting in 100% adherence to the tDCS protocol.

### 3.1. Active tDCS

**Trajectory Model Results (Table 1 and Figure 2)**. All fit criteria favored models with a greater number of classes. However, we selected the two-group multi-trajectory model (AIC = -5312.39, BIC = - 5342.76) for several reasons. Although the three-class model demonstrated slightly better fit statistics than the two-class model, it merely added a class reflecting the average levels of the other two without revealing substantively distinct trajectory patterns. Similarly, the four-class model included a marginally distinct class with a relatively small number of participants, making it less interpretable. Consequently, the two-class model most effectively captured substantially differentiated and meaningful trajectory patterns. Group 1 comprised 46.9% of participants (*n* = 28), while Group 2 included the remaining 53.1% (*n* = 31).

**Figure 2.**
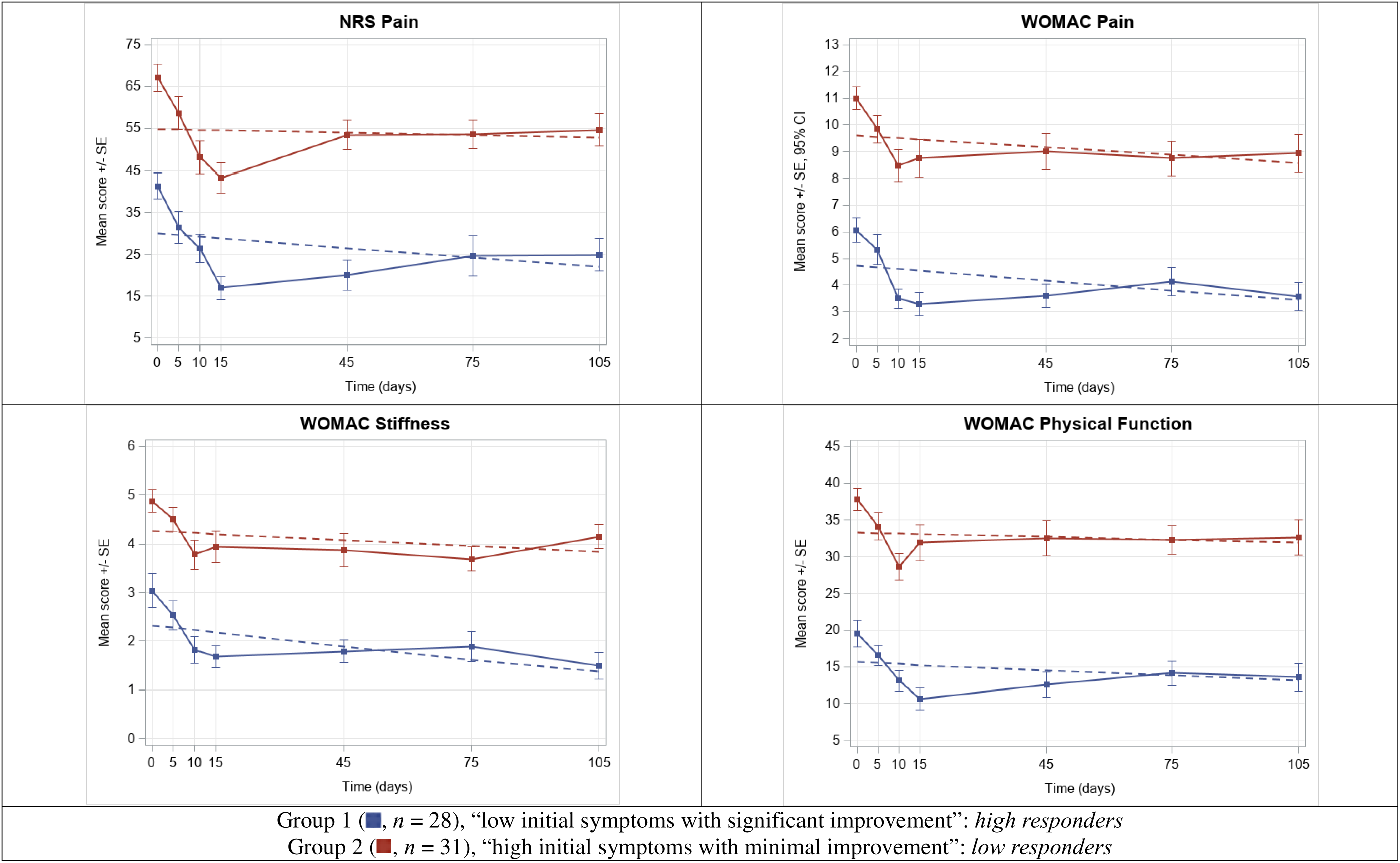
Two-group multi-trajectory model: active tDCS. *Note.* The dots represent the mean observed levels, while the solid line represents the expected trajectories. The error bars denote the standard error (SE). Please see **Supplpplemental Table 3** for the descriptive statistics of the mean observed levels presented in the graph. *Abbreviation.* NRS, Numeric Rating Scale; tDCS, transcranial direct current stimulation; WOMAC, Western Ontario and McMaster Universities Osteoarthritis Index

**Table 1.** Two-group multi-trajectory model results.

| | Engagement group | Parameter | $\beta$ (estimate) | Standard error | *t*-value | *p*-value |
| --- | --- | --- | --- | --- | --- | --- |
| *Active tDCS* |  |  |  |  |  |  |
| Group membership (%) | Group 1 |  | 46.91 | 6.55 | 7.17 | <.000 |
|  | Group 2 |  | 53.09 | 6.55 | 8.11 | <.000 |
| NRS pain | Group 1 | linear | -0.09 | 0.04 | -2.08 | **.037** |
|  | Group 2 | linear | -0.02 | 0.04 | -0.53 | .599 |
| WOMAC pain | Group 1 | linear | -0.01 | 0.01 | -2.10 | **.036** |
|  | Group 2 | linear | -0.01 | 0.01 | -1.59 | .113 |
| WOMAC stiffness | Group 1 | linear | -0.01 | 0.003 | -3.20 | **.001** |
|  | Group 2 | linear | -0.004 | 0.003 | -1.33 | .182 |
| WOMAC physical function | Group 1 | linear | -0.03 | 0.02 | -1.27 | .205 |
|  | Group 2 | linear | -0.47 | 0.38 | -1.25 | .212 |
| *Sham tDCS* |  |  |  |  |  |  |
| Group membership (%) | Group 1 |  | 55.40 | 6.56 | 8.44 | <.000 |
|  | Group 2 |  | 44.60 | 6.56 | 6.80 | <.000 |
| NRS pain | Group 1 | linear | -0.002 | 0.04 | -0.05 | .962 |
|  | Group 2 | linear | 0.05 | 0.04 | 1.25 | .213 |
| WOMAC pain | Group 1 | linear | -0.01 | 0.005 | -2.94 | **.003** |
|  | Group 2 | linear | -0.01 | 0.005 | 2.63 | **.009** |
| WOMAC stiffness | Group 1 | linear | -0.01 | 0.003 | -2.35 | **.019** |
|  | Group 2 | linear | -0.01 | 0.003 | 2.97 | **.003** |
| WOMAC physical function | Group 1 | linear | -0.02 | 0.02 | -1.50 | .133 |
|  | Group 2 | linear | -0.03 | 0.02 | -1.84 | .066 |

*Note.* Active tDCS: Akaike Information Criteria (AIC) = -5320.22, Bayesian Information Criteria (BIC) = -5377.20
Sham tDCS: Akaike Information Criteria (AIC) = -5185.25, Bayesian Information Criteria (BIC) = -5242.23
Statistically significant *p*-values were written in **bold**.
*Abbreviation.* NRS, Numeric Rating Scale; tDCS, transcranial direct current stimulation; WOMAC, Western Ontario and McMaster Universities Osteoarthritis Index

Group 2 exhibited higher baseline levels for KOA pain and symptoms compared to Group 1. Both groups demonstrated an overall linear decrease in symptoms from baseline to the three-month follow-up, with all measures generally declining over the 15-day period (+15 active tDCS sessions), despite slight increases during the monthly follow-ups. However, these changes were not statistically significant in Group 2 (NRS: β = -0.02, *p* = .599; WOMAC pain: β = -0.01, *p* = .113; WOMAC stiffness: β = -0.004, *p* = .182; WOMAC physical function: β = -0.01, *p* = .518). In contrast, Group 1 showed significant linear reductions in most symptoms, particularly in pain NRS (NRS: β = -0.09, *p* = .037; WOMAC pain: β = - 0.01, *p* = .036; and WOMAC stiffness: β = -0.01, *p* = .001) throughout the study. Based on these findings, Group 1 was designated the “low initial symptoms with significant improvement” group or “high responders,” whereas Group 2 was designated the “high initial symptoms with minimal improvement” group or “low responders.”

**Characteristics of the Participants by Trajectory Group (Table 2).** Group 2 had a significantly higher BMI than Group 1 (*p* = .040), with BMI values of 34.93 ± 10.00 kg/m² and 30.09 ± 6.23 kg/m², respectively. Additionally, a greater proportion of participants in Group 1 had at least a two-year college degree compared to Group 2 (*p* = .013), with nearly 93% of Group 1 achieving this level of education or higher. Group 2 exhibited significantly greater pain catastrophizing levels than Group 1, with PCS scores of 22.16 ± 14.73 and 8.21 ± 8.45, respectively (*p* < .000). Regarding QST measures, significant baseline differences were observed between the groups in PPTh at the medial knee (*p* = .009) and trapezius (*p* = .002), punctate mechanical pain at the patella (*p* = .013) and hand (*p* = .016), CPM at 30 seconds (*p* = .008) and 60 seconds (*p* < .000), and cold pain intensity at 30 seconds (*p* = .003). Compared to Group 1, Group 2 demonstrated lower PPTh at both sites, higher punctate mechanical pain at both sites, lower CPM, and higher cold pain intensity. No significant differences were found between the groups (*p* > .05) in age, gender, race, education, marital status, Kellgren-Lawrence score, average duration of osteoarthritis, HPTh, HPTo, or TSP.

**Table 2.**
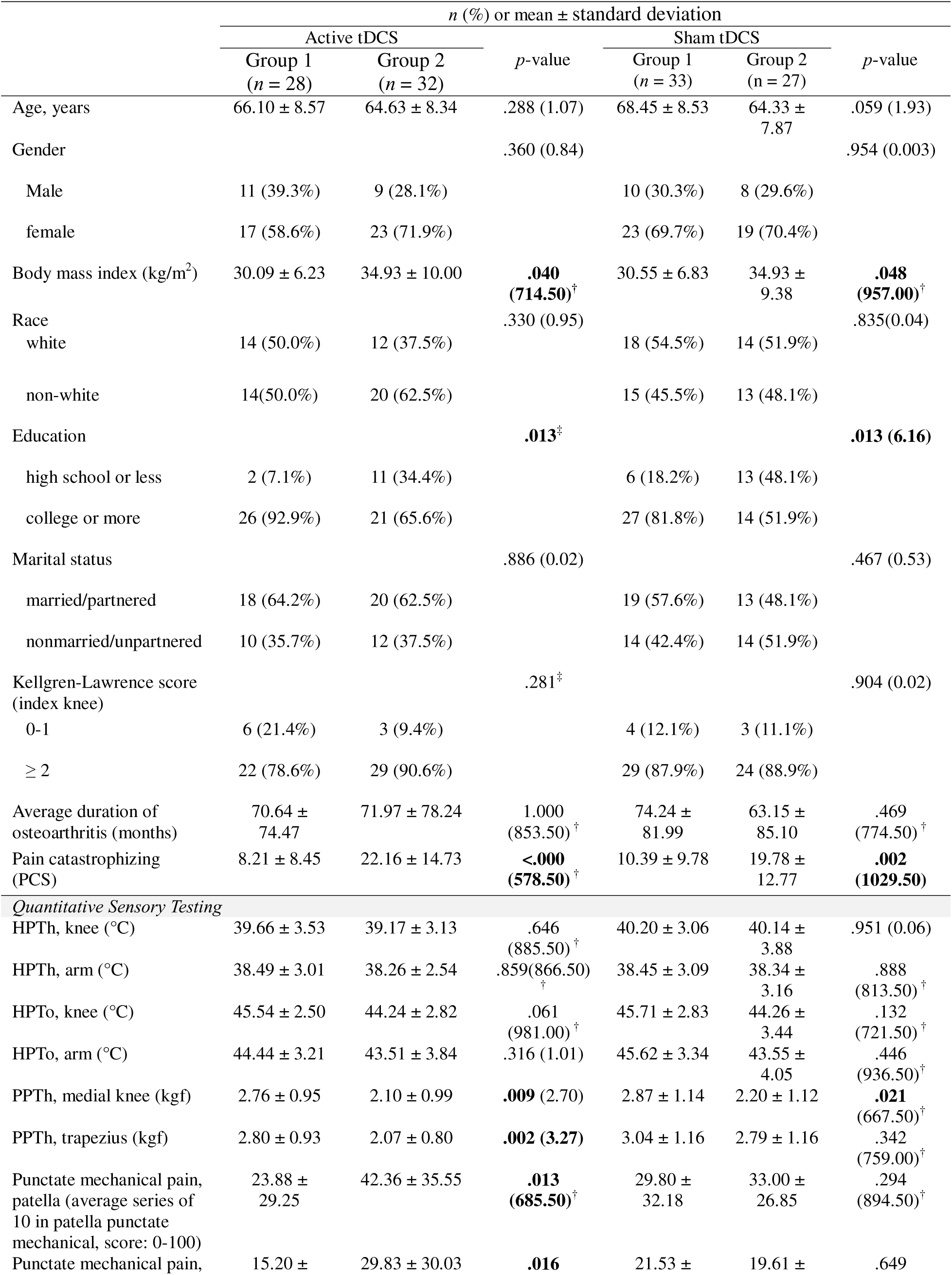
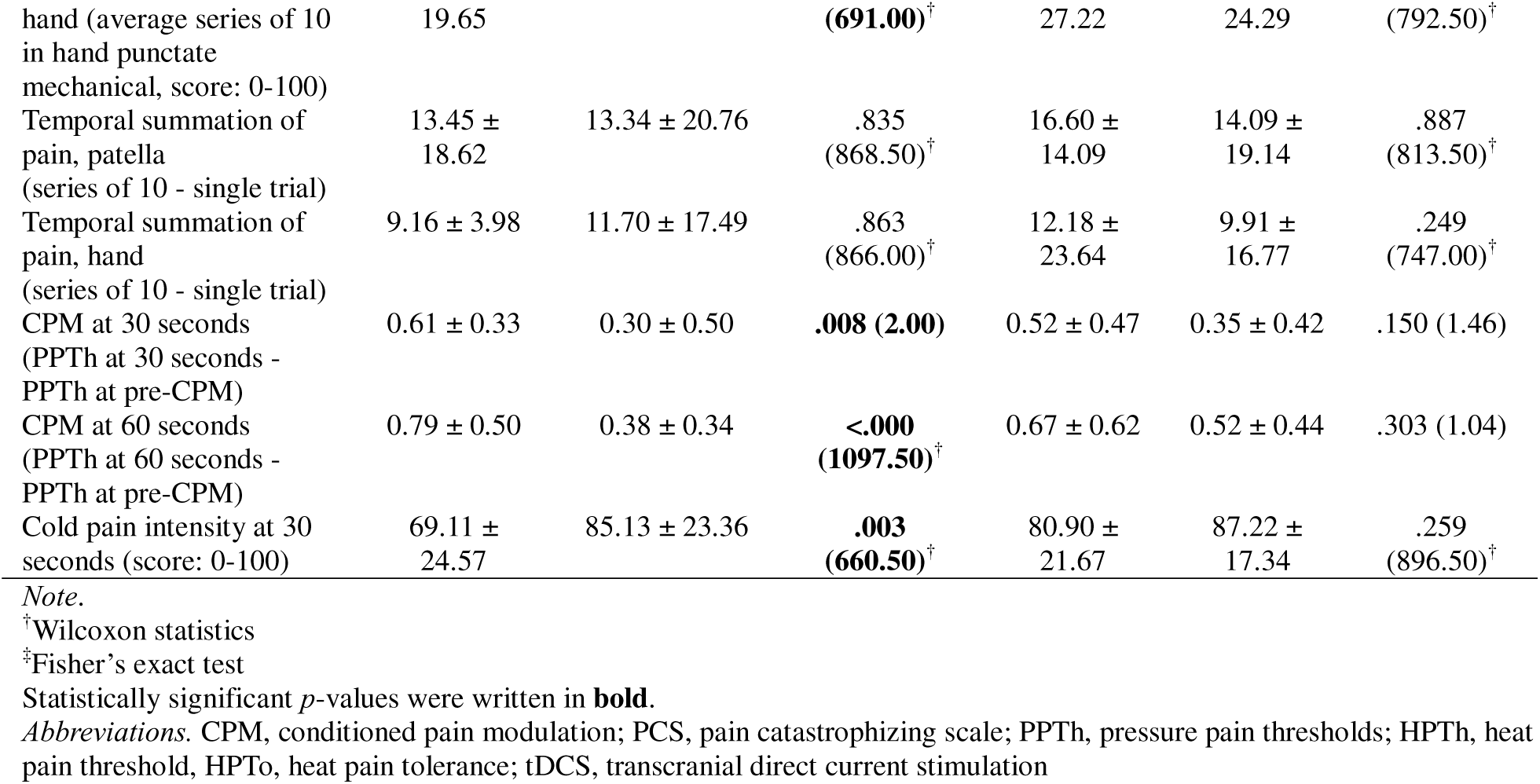
Patient characteristics within each group.

### 3.1. Sham tDCS

**Trajectory Model Results (Table 1 and Figure 3)** The two-group multi-trajectory model was selected as the final model based on the distinctiveness of the groups and their clinical relevance. The three-class model did not yield any meaningful additional insights, while the four-class model was more complex and less interpretable compared to the more parsimonious two-class solution. Group 1 comprised 55.4% of participants (*n* = 33), while the remaining 44.6% were in Group 2 (*n* = 27).

**Figure 3.**
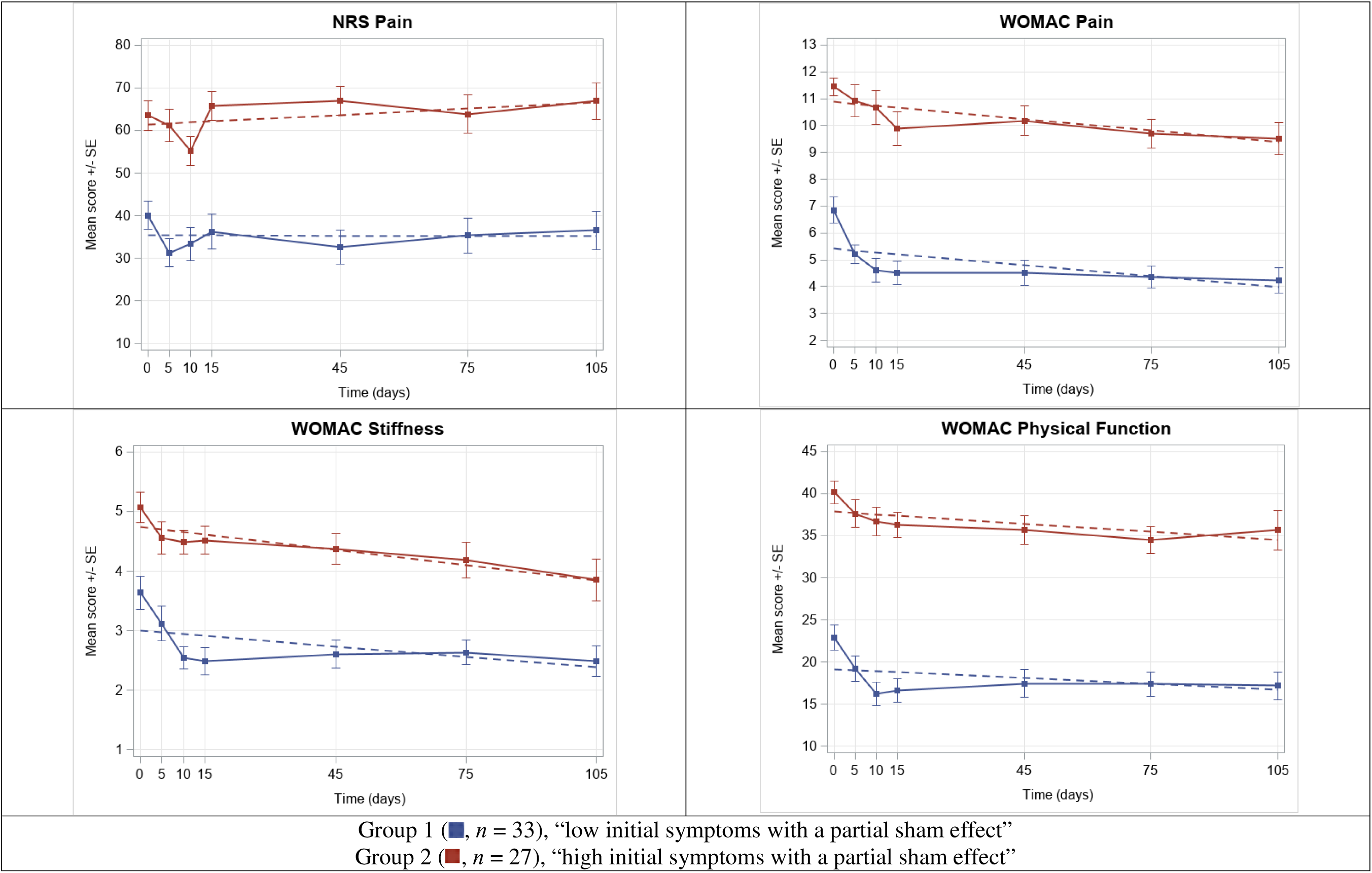
**Two-group multi-trajectory model: sham tDCS** *Note.* The dots represent the mean observed levels, while the solid line represents the expected trajectories. The error bars denote the standard error (SE). Please see **Supplpplemental Table 4** for the descriptive statistics of the mean observed levels presented in the graph. *Abbreviation.* NRS, Numeric Rating Scale; tDCS, transcranial direct current stimulation; WOMAC, Western Ontario and McMaster Universities Osteoarthritis Index

Group 2 exhibited higher baseline values for all KOA pain and symptom measures compared to Group 1. Throughout the study, significant linear decreases were observed in WOMAC pain and stiffness in both groups (all *p* < .050), with symptom reductions occurring over the 15-day period (+15 sham tDCS sessions). In Group 2, NRS pain showed an increasing trend; however, these changes were not statistically significant (β = 0.05, *p* = .213). Group 1 was designated as “low initial symptoms with a partial sham effect,” while Group 2 was classified as “high initial symptoms with a partial sham effect.”

**Characteristics of the Participants by Trajectory Group (Table 2)** Group 2 had a higher BMI than Group 1 (*p* = .048), with values of 34.93 ± 9.38 kg/m² and 30.55 ± 6.83 kg/m², respectively. A greater proportion of participants in Group 1 had completed at least a two-year college degree compared to Group 2 (*p* = .013), with nearly 82% of individuals in Group 1 achieving this level of education or higher. Group 2 exhibited significantly higher pain catastrophizing than Group 1 (*p* = .002), with PCS scores of 19.78 ± 12.77 in Group 2 and 10.39 ± 9.78 in Group 1. Regarding QST measures, a significant baseline difference was observed in PPTh at the medial knee (*p* = .021), with Group 2 demonstrating lower PPTh than Group 1 (Group 1: 2.87 ± 1.14 kgf; Group 2: 2.20 ± 1.12 kgf). No significant baseline differences were found between the groups for other factors.

## 4. Discussion

The study identified HTE of tDCS in participants with symptomatic KOA, as well as key participant characteristics associated with this heterogeneity. In the active tDCS group, two distinct response trajectories emerged: “low initial symptoms with significant improvement” and “high initial symptoms with minimal improvement.” The former group can be classified as “high responders,” while the latter represents “low responders.” Several baseline characteristics distinguished high responders from low responders, including BMI, education, pain catastrophizing, PPTh at both the medial knee and trapezius, punctate mechanical pain at both the patella and hand, cold pain, and CPM.

In this study, low responders had a higher BMI than high responders. The precise mechanism underlying this relationship remains unclear; however, comorbid health conditions associated with KOA may exacerbate pain and functional limitations, potentially hindering an individual’s ability to fully engage in the intervention. Additionally, evidence suggests that obesity is linked to structural and functional brain alterations, as well as cognitive deficits, which may stem from altered neuroplasticity and could contribute to an impaired response to brain stimulation.^32^ Furthermore, biomechanical factors, such as increased joint loading, may influence the analgesic response to tDCS.

Low responders had lower educational attainment than high responders. Previous tDCS studies have shown that education moderates its effects on memory performance and cognitive outcomes, with more years of education associated with greater benefits—underscoring the role of education in personalized stimulation protocols.^33–35^ Similarly, de Rooij et al.^12^ found that higher educational attainment contributed to greater improvements in pain interference among individuals undergoing multidisciplinary treatment for chronic widespread pain. The present findings may be attributed to differences in health literacy, which could influence participants’ comprehension of tDCS-related materials and adherence to stimulation procedures. Even with remote clinical supervision, these factors may have impacted the response to tDCS.

Low responders exhibit higher levels of pain catastrophizing at baseline compared to high responders. This finding is consistent with previous research indicating that pain catastrophizing is one of the strongest predictors of pain treatment outcomes across various chronic pain conditions, including chronic neuropathic pain, musculoskeletal pain, low back pain, and fibromyalgia.^36–39^ According to the biopsychosocial model, pain perception and response are shaped by the complex interplay of physiological, psychological, and social factors.^40^ For individuals with high pain catastrophizing, addressing this factor should be a key consideration in future tDCS trials. One potential approach is to combine tDCS with other active interventions that specifically target pain catastrophizing, such as cognitive behavioral therapy.^41^ Additionally, neurofunctional imaging studies suggest that pain catastrophizing is associated with increased activity in the dorsolateral prefrontal cortex (DLPFC).^42^ The DLPFC plays a significant role in the cognitive processing of pain, particularly in pain prediction, evaluation, and reinterpretation.^42,43^ The left DLPFC (F3) is commonly targeted in tDCS research due to its critical role in cognitive functions and emotion regulation. Previous studies have demonstrated that F3 stimulation reduces pain catastrophizing.^44,45^

Low responders exhibited greater sensitivity to pressure, mechanical pain (at both the affected knee and remote body sites), and cold pain compared to high responders. Increased hypersensitivity at the affected site suggests peripheral sensitization, while hypersensitivity at an unaffected site indicates central sensitization.^46^ Although no prior tDCS studies have specifically examined this relationship, a systematic review and meta-analysis found that pretreatment QST profiles indicative of both peripheral and central sensitization are associated with reduced pain relief following total knee arthroplasty, nonsteroidal anti-inflammatory drug (NSAID) treatment, and exercise-based therapies.^47^ Peripheral pain mechanisms in KOA primarily originate in the joint structures, particularly the synovium and subchondral bone.^48^ Neogi et al.^49^ reported that lower PPTh at the affected joint in KOA correlates with the degree of synovitis observed on functional magnetic resonance imaging (MRI). Although the present study did not assess structural joint changes via MRI, joint inflammation and other pathological factors may have contributed to the limited reduction in KOA pain and symptoms among study participants. In case of central sensitization, various alterations in somatosensory input processing mechanisms and central nervous system neurons become overstimulated, and this heightened pain sensitization may contribute to a diminished response to pain treatment. In individuals with KOA experiencing central sensitization, clinicians should consider whether stimulation parameters of tDCS (e.g., intensity, duration, session frequency, and inter-session intervals) must be adjusted and tailored accordingly. Furthermore, future tDCS trials could benefit from personalized approaches that account for pain sensitization, such as pain neuroscience education,^50^ to optimize treatment outcomes in affected individuals.

Additionally, low responders exhibited lower CPM than high responders, suggesting that a less efficient endogenous pain inhibitory system may contribute to an inadequate response to tDCS. The previously mentioned systematic review and meta-analysis identified CPM as the second most frequently reported QST parameter (40%) for pharmacological treatment following temporal stimulation of pain (TSP; 60%).^47^ However, no prior studies have specifically examined the relationship between CPM and responses to tDCS.

No HTE was observed in the sham tDCS group; both trajectory groups demonstrated a partial placebo response, characterized by reductions in WOMAC pain and stiffness, but differed in their initial symptom severity—categorized as low or high. The findings on the partial sham effect suggest that a sham protocol previously considered inactive may still exert minor neuromodulatory effects on certain outcomes.^51^ This aligns with prior studies,^52,53^ which suggest that such effects may arise from skin sensations initially induced in the sham condition (e.g., ramp-up/down procedures) or cortical modulation by micro-ampere-scale currents. However, caution is warranted when interpreting these findings, as the observed symptom changes may also reflect the natural course of pain rather than a direct neuromodulatory effect.

Several limitations should be acknowledged. First, these analyses were exploratory and not prespecified in the trial, necessitating cautious interpretation of the conclusions. Second, the modest sample size resulted in small subgroup sizes, limiting the generalizability of the findings to broader KOA populations. Third, the study does not establish causal relationships but instead generates hypotheses regarding potential mechanisms underlying the observed HTE. Fourth, the observed baseline differences between trajectory groups were, at least partly, influenced by the limited set of baseline variables included in our analysis. Future studies should consider incorporating a broader range of participant characteristics for a more comprehensive characterization of trajectory group differences. Fifth, treatment decisions based on the group-level analyses may be suboptimal, as they rely on individual participant characteristics in isolation, whereas a combination of factors likely influences the treatment response, which should be further explored.

## 5. Conclusion

Participants with symptomatic KOA exhibited varying responses to tDCS. The characteristics associated with HTE, which distinguished high responders from low responders in this exploratory study, provide preliminary insights that may influence the development of personalized stimulation protocols. Continued efforts in this area could ultimately improve the efficacy and applicability of neuromodulatory interventions for chronic pain and symptom management in KOA, potentially informing the development of more appropriate brain stimulation models. Additionally, further research is needed to explore potential HTE in sham tDCS within sham-controlled tDCS trials. Additionally, further research is warranted to explore potential HTE in sham tDCS within sham-controlled tDCS trials.

## Data Availability

All data produced in the present study are available upon reasonable request to the authors.

## References

1. Courties A, Kouki I, Soliman N, Mathieu S, Sellam J. Osteoarthritis year in review 2024: Epidemiology and therapy. Osteoarthritis Cartilage 2024; 32(11): 1397–404.

2. Deshpande BR, Katz JN, Solomon DH, et al. Number of Persons With Symptomatic Knee Osteoarthritis in the US: Impact of Race and Ethnicity, Age, Sex, and Obesity. Arthritis Care Res (Hoboken) 2016; 68(12): 1743–50.

3. Hawker GA, King LK. The Burden of Osteoarthritis in Older Adults. Clin Geriatr Med 2022; 38(2): 181–92.

4. Clauw DJ, Hassett AL. The role of centralised pain in osteoarthritis. Clin Exp Rheumatol 2017; 35 Suppl 107(5): 79–84.

5. Lluch E, Nijs J, Courtney CA, et al. Clinical descriptors for the recognition of central sensitization pain in patients with knee osteoarthritis. Disabil Rehabil 2018; 40(23): 2836–45.

6. Pacheco-Barrios K, Cardenas-Rojas A, Thibaut A, et al. Methods and strategies of tDCS for the treatment of pain: current status and future directions. Expert Rev Med Devices 2020; 17(9): 879–98.

7. Ahn H, Woods AJ, Kunik ME, et al. Efficacy of transcranial direct current stimulation over primary motor cortex (anode) and contralateral supraorbital area (cathode) on clinical pain severity and mobility performance in persons with knee osteoarthritis: An experimenter- and participant-blinded, randomized, sham-controlled pilot clinical study. Brain Stimul 2017; 10(5): 902–9.

8. Ahn H, Suchting R, Woods AJ, et al. Bayesian analysis of the effect of transcranial direct current stimulation on experimental pain sensitivity in older adults with knee osteoarthritis: randomized sham-controlled pilot clinical study. J Pain Res 2018; 11: 2071–82.

9. Ahn H, Sorkpor S, Miao H, et al. Home-based self-administered transcranial direct current stimulation in older adults with knee osteoarthritis pain: An open-label study. J Clin Neurosci 2019; 66: 61–5.

10. Martorella G, Mathis K, Miao H, Wang D, Park L, Ahn H. Efficacy of Home-Based Transcranial Direct Current Stimulation on Experimental Pain Sensitivity in Older Adults with Knee Osteoarthritis: A Randomized, Sham-Controlled Clinical Trial. J Clin Med 2022; 11(17).

11. Kaplan SH, Billimek J, Sorkin DH, Ngo-Metzger Q, Greenfield S. Who can respond to treatment? Identifying patient characteristics related to heterogeneity of treatment effects. Med Care 2010; 48(6 Suppl): S9–16.

12. de Rooij A, van der Leeden M, Roorda LD, Steultjens MP, Dekker J. Predictors of outcome of multidisciplinary treatment in chronic widespread pain: an observational study. BMC Musculoskelet Disord 2013; 14: 133.

13. Boonstra AM, Reneman MF, Waaksma BR, Schiphorst Preuper HR, Stewart RE. Predictors of multidisciplinary treatment outcome in patients with chronic musculoskeletal pain. Disabil Rehabil 2015; 37(14): 1242–50.

14. Willke RJ, Zheng Z, Subedi P, Althin R, Mullins CD. From concepts, theory, and evidence of heterogeneity of treatment effects to methodological approaches: a primer. BMC Med Res Methodol 2012; 12: 185.

15. Knotkova H, Riggs A, Berisha D, et al. Automatic M1-SO Montage Headgear for Transcranial Direct Current Stimulation (TDCS) Suitable for Home and High-Throughput In-Clinic Applications. Neuromodulation 2019; 22(8): 904–10.

16. DosSantos MF, Ferreira N, Toback RL, Carvalho AC, DaSilva AF. Potential Mechanisms Supporting the Value of Motor Cortex Stimulation to Treat Chronic Pain Syndromes. Front Neurosci 2016; 10: 18.

17. Lefaucheur JP, Antal A, Ayache SS, et al. Evidence-based guidelines on the therapeutic use of transcranial direct current stimulation (tDCS). Clin Neurophysiol 2017; 128(1): 56–92.

18. Mendonca ME, Santana MB, Baptista AF, et al. Transcranial DC stimulation in fibromyalgia: optimized cortical target supported by high-resolution computational models. J Pain 2011; 12(5): 610–7.

19. Hawker GA, Mian S, Kendzerska T, French M. Measures of adult pain: Visual Analog Scale for Pain (VAS Pain), Numeric Rating Scale for Pain (NRS Pain), McGill Pain Questionnaire (MPQ), Short-Form McGill Pain Questionnaire (SF-MPQ), Chronic Pain Grade Scale (CPGS), Short Form-36 Bodily Pain Scale (SF-36 BPS), and Measure of Intermittent and Constant Osteoarthritis Pain (ICOAP). Arthritis Care Res (Hoboken) 2011; 63 Suppl 11: S240–52.

20. Bellamy N, Buchanan WW, Goldsmith CH, Campbell J, Stitt LW. Validation study of WOMAC: a health status instrument for measuring clinically important patient relevant outcomes to antirheumatic drug therapy in patients with osteoarthritis of the hip or knee. J Rheumatol 1988; 15(12): 1833–40.

21. Bellamy N, Kirwan J, Boers M, et al. Recommendations for a core set of outcome measures for future phase III clinical trials in knee, hip, and hand osteoarthritis. Consensus development at OMERACT III. J Rheumatol 1997; 24(4): 799–802.

22. Sullivan MJ, Stanish WD. Psychologically based occupational rehabilitation: the Pain-Disability Prevention Program. Clin J Pain 2003; 19(2): 97–104.

23. Sullivan MJ, Thorn B, Haythornthwaite JA, et al. Theoretical perspectives on the relation between catastrophizing and pain. Clin J Pain 2001; 17(1): 52–64.

24. Sullivan MJ, Ward LC, Tripp D, French DJ, Adams H, Stanish WD. Secondary prevention of work disability: community-based psychosocial intervention for musculoskeletal disorders. J Occup Rehabil 2005; 15(3): 377–92.

25. Arendt-Nielsen L, Yarnitsky D. Experimental and clinical applications of quantitative sensory testing applied to skin, muscles and viscera. J Pain 2009; 10(6): 556–72.

26. Uddin Z, MacDermid JC. Quantitative Sensory Testing in Chronic Musculoskeletal Pain. Pain Med 2016; 17(9): 1694–703.

27. Arendt-Nielsen L, Morlion B, Perrot S, et al. Assessment and manifestation of central sensitisation across different chronic pain conditions. Eur J Pain 2018; 22(2): 216–41.

28. Yarnitsky D, Arendt-Nielsen L, Bouhassira D, et al. Recommendations on terminology and practice of psychophysical DNIC testing. Eur J Pain 2010; 14(4): 339.

29. King CD, Sibille KT, Goodin BR, et al. Experimental pain sensitivity differs as a function of clinical pain severity in symptomatic knee osteoarthritis. Osteoarthritis Cartilage 2013; 21(9): 1243–52.

30. Nagin DS, Jones BL, Passos VL, Tremblay RE. Group-based multi-trajectory modeling. Stat Methods Med Res 2018; 27(7): 2015–23.

31. Muthen B, Muthen LK. Integrating person-centered and variable-centered analyses: growth mixture modeling with latent trajectory classes. Alcohol Clin Exp Res 2000; 24(6): 882–91.

32. Sui SX, Ridding MC, Hordacre B. Obesity is Associated with Reduced Plasticity of the Human Motor Cortex. Brain Sci 2020; 10(9).

33. Berryhill ME, Jones KT. tDCS selectively improves working memory in older adults with more education. Neurosci Lett 2012; 521(2): 148–51.

34. Krebs C, Kloppel S, Heimbach B, Peter J. Education moderates the effect of tDCS on episodic memory performance in cognitively impaired patients. Brain Stimul 2020; 13(5): 1396–8.

35. Krebs C, Peter J, Brill E, Kloppel S, Brem AK. The moderating effects of sex, age, and education on the outcome of combined cognitive training and transcranial electrical stimulation in older adults. Front Psychol 2023; 14: 1243099.

36. Domenech J, Sanchis-Alfonso V, Espejo B. Changes in catastrophizing and kinesiophobia are predictive of changes in disability and pain after treatment in patients with anterior knee pain. Knee Surg Sports Traumatol Arthrosc 2014; 22(10): 2295–300.

37. Farin E. The reciprocal effect of pain catastrophizing and satisfaction with participation in the multidisciplinary treatment of patients with chronic back pain. Health Qual Life Outcomes 2015; 13: 163.

38. Oosterhaven J, Wittink H, Dekker J, Kruitwagen C, Deville W. Pain catastrophizing predicts dropout of patients from an interdisciplinary chronic pain management programme: A prospective cohort study. J Rehabil Med 2019; 51(10): 761–9.

39. Shaygan M, Boger A, Kroner-Herwig B. Predicting factors of outcome in multidisciplinary treatment of chronic neuropathic pain. J Pain Res 2018; 11: 2433–43.

40. Gatchel RJ. Comorbidity of chronic pain and mental health disorders: the biopsychosocial perspective. Am Psychol 2004; 59(8): 795–805.

41. Burns JW, Day MA, Thorn BE. Is reduction in pain catastrophizing a therapeutic mechanism specific to cognitive-behavioral therapy for chronic pain? Transl Behav Med 2012; 2(1): 22–9.

42. Galambos A, Szabo E, Nagy Z, et al. A systematic review of structural and functional MRI studies on pain catastrophizing. J Pain Res 2019; 12: 1155–78.

43. Arul-Anandam AP, Loo C, Martin D, Mitchell PB. Chronic neuropathic pain alleviation after transcranial direct current stimulation to the dorsolateral prefrontal cortex. Brain Stimul 2009; 2(3): 149–51.

44. Caumo W, Alves RL, Vicuna P, et al. Impact of Bifrontal Home-Based Transcranial Direct Current Stimulation in Pain Catastrophizing and Disability due to Pain in Fibromyalgia: A Randomized, Double-Blind Sham-Controlled Study. J Pain 2022; 23(4): 641–56.

45. Caumo W, Lopes Ramos R, Vicuna Serrano P, et al. Efficacy of Home-Based Transcranial Direct Current Stimulation Over the Primary Motor Cortex and Dorsolateral Prefrontal Cortex in the Disability Due to Pain in Fibromyalgia: A Factorial Sham-Randomized Clinical Study. J Pain 2024; 25(2): 376–92.

46. Ohashi Y, Uchida K, Fukushima K, Inoue G, Takaso M. Mechanisms of Peripheral and Central Sensitization in Osteoarthritis Pain. Cureus 2023; 15(2): e35331.

47. Petersen KK, Kilic K, Hertel E, et al. Quantitative sensory testing as an assessment tool to predict the response to standard pain treatment in knee osteoarthritis: a systematic review and meta-analysis. Pain Rep 2023; 8(4): e1079.

48. Vincent TL. Peripheral pain mechanisms in osteoarthritis. Pain 2020; 161 Suppl 1(1): S138–S46.

49. Neogi T, Guermazi A, Roemer F, et al. Association of Joint Inflammation With Pain Sensitization in Knee Osteoarthritis: The Multicenter Osteoarthritis Study. Arthritis Rheumatol 2016; 68(3): 654–61.

50. Lepri B, Romani D, Storari L, Barbari V. Effectiveness of Pain Neuroscience Education in Patients with Chronic Musculoskeletal Pain and Central Sensitization: A Systematic Review. Int J Environ Res Public Health 2023; 20(5).

51. Braga M, Barbiani D, Emadi Andani M, Villa-Sanchez B, Tinazzi M, Fiorio M. The Role of Expectation and Beliefs on the Effects of Non-Invasive Brain Stimulation. Brain Sci 2021; 11(11).

52. Creutzfeldt OD, Fromm GH, Kapp H. Influence of transcortical d-c currents on cortical neuronal activity. Exp Neurol 1962; 5: 436–52.

53. Nikolin S, Martin D, Loo CK, Boonstra TW. Effects of TDCS dosage on working memory in healthy participants. Brain Stimul 2018; 11(3): 518–27.

