## Supplemental Tables for "Part 1: Examining heterogeneity of treatment effects in transcranial direct current stimulation for knee osteoarthritis pain and symptoms"

### **Supplemental Table 1.** Baseline characteristics of each treatment group (*n* =120)

|  | *n* (%) or mean ± standard deviation | |
| --- | --- | --- |
|  | Active tDCS (*n* = 60) | Sham tDCS (*n* = 60) |
| Age, years | 65.32 ± 8.41 | 66.60 ± 8.43 |
| Gender |  |  |
| Male | 20 (33.3%) | 18 (30.0%) |
| female | 60 (60.7%) | 42 (70.0%) |
| Body mass index (kg/m^2^) | 32.67 ± 8.73 | 32.52 ± 8.30 |
| Race |  |  |
| white | 26 (43.3%) | 32 (53.3%) |
| non-white | 34 (56.7%) | 28 (46.7%) |
| Education |  |  |
| high school or less | 13 (21.7%) | 19 (31.7%) |
| college or more | 47 (78.3%) | 41 (68.3%) |
| Marital status |  |  |
| married/partnered | 38 (63.3%) | 32 (53.3%) |
| nonmarried/unpartnered | 22 (36.7%) | 28 (46.7%) |
| Kellgren-Lawrence score (Index knee) |  |  |
| 0-1 | 9 (15.0%) | 7 (11.7%) |
| ≥ 2 | 51 (85.0%) | 53 (88.3%) |
| Average duration of osteoarthritis (months) | 71.35 ± 75.23 | 69.25 ± 82.18 |
| Pain catastrophizing (PCS) | 15.65 ± 13.99 | 14.62 ± 12.08 |
| *Quantitative sensory testing* |  |  |
| HPTh, knee (°C) | 39.40 ± 3.30 | 40.17 ± 3.42 |
| HPTh, arm (°C) | 38.37 ± 2.75 | 38.40 ± 3.10 |
| HPTo, knee (°C) | 44.85 ± 2.74 | 45.06 ± 3.17 |
| HPTo, arm (°C) | 43.94 ± 3.56 | 44.69 ± 3.79 |
| PPTh, medial knee (kgf) | 2.40 ± 1.02 | 2.57 ± 1.17 |
| PPTh, trapezius (kgf) | 2.41 ± 0.93 | 2.93 ± 1.16 |
| Punctate mechanical pain, patella (average series of 10 in patella punctate mechanical, score: 0-100) | 33.73 ± 33.79 | 31.24 ± 29.70 |
| Punctate mechanical pain, hand (average series of 10 in patella punctate mechanical, score: 0-100) | 26.55 ± 1.69 | 20.67 ± 25.75 |
| Temporal summation of pain, patella  (series of 10 - single trial) | 13.39 ± 19.62 | 15.46 ± 21.76 |
| Temporal summation of pain, hand  (series of 10 - single trial) | 10.52 ± 15.62 | 11.16 ± 20.70 |
| CPM at 30 seconds  (PPTh at 30 seconds - PPTh at pre-CPM) | 0.44 ± 0.45 | 0.45 ± 0.45 |
| CPM at 60 seconds  (PPTh at 60 seconds - PPTh at pre-CPM) | 0.60 ± 0.47 | 0.60 ± 0.55 |
| Cold pain intensity at 30 seconds (score: 0-100) | 77.65 ± 25.06 | 83.75 ± 19.93 |

*Note*. CPM, conditioned pain modulation; PCS, pain catastrophizing scale; PPTh, pressure pain thresholds; HPTh, heat pain threshold, HPTo, heat pain tolerance; tDCS, transcranial direct current stimulation

### **Supplemental Table 2.** Fit statistics for multi-trajectory models

| No. of classes | % per class | AIC | BIC |
| --- | --- | --- | --- |
| *Active tDCS* | | | |
| 2 (linear) | 46.9%, 53.1% | -5320.22 | -5377.20 |
| 2 (quadratic) | 48.7%, 51.3% | -5312.39 | -5391.07 |
| 3 (linear) | 28.4%, 46.6%, 25.0% | -5163.79 | -5245.19 |
| 3 (quadratic) | 28.4%, 46.5%, 25.1% | -5145.86 | -5259.82 |
| 4 (linear) | 24.3%, 17.5%, 32.8%, 23.4% | -5129.63 | -5235.44 |
| 4 (quadratic) | 23.8%, 13.9%, 37.3%, 25.0% | -5112.64 | -5261.87 |
| *Sham tDCS* | | | |
| 2 (linear) | 55.4%, 44.6% | -5185.25 | -5242.23 |
| 2 (quadratic) | 55.7%, 44.3% | -5186.85 | -5265.54 |
| 3 (linear) | 26.7%, 40.0%, 33.3% | -5059.99 | -5141.39 |
| 3 (quadratic) | 26.8%, 39.9%, 33.3% | -5064.31 | -5178.27 |
| 4 (linear) | 13.2%, 38.4%, 27.9%, 20.5% | -5020.31 | -5126.12 |
| 4 (quadratic) | 11.9%, 39.3%, 18.9%, 29.9% | -5020.12 | -5169.35 |

*Abbreviations.* AIC, Akaike Information Criteria; BIC, Bayesian Information Criteria; tDCS, transcranial direct current stimulation

### **Supplemental Table 3.** Descriptive statistics for all measures throughout the study: active tDCS

|  |  | mean ± standard deviation | |
| --- | --- | --- | --- |
|  | All | Group 1 | Group 2 |
| *NRS pain* |  |  |  |
| Baseline | 55.05 ± 21.96 | 41.25 ± 16.48 | 67.13 ± 18.95 |
| 5 days | 45.93 ± 24.94 | 31.39 ± 19.73 | 58.66 ± 22.06 |
| 10 days | 37.98 ± 22.84 | 26.39 ± 18.25 | 48.13 ± 21.79 |
| 15 days | 30.98 ± 22.21 | 16.96 ± 14.44 | 43.25 ± 20.59 |
| 45 days | 37.85 ± 25.64 | 20.04 ± 19.06 | 54.44 ± 19.94 |
| 75 days | 40.08 ± 26.43 | 24.57 ± 25.13 | 53.66 ± 19.29 |
| 105 days | 40.78 ± 26.01 | 24.89 ± 20.94 | 54.69 ± 21.85 |
| *WOMAC pain* |  |  |  |
| Baseline | 8.70 ± 3.44 | 6.07 ± 2.46 | 11.00 ± 2.34 |
| 5 days | 7.73 ± 3.70 | 5.32 ± 3.01 | 9.84 ± 2.90 |
| 10 days | 6.15 ± 3.74 | 3.50 ± 1.90 | 8.47 ± 3.40 |
| 15 days | 6.20 ± 4.28 | 3.29 ± 2.34 | 8.75 ± 3.96 |
| 45 days | 6.48 ± 4.20 | 3.61 ± 2.33 | 9.00 ± 3.84 |
| 75 days | 6.60 ± 4.00 | 4.14 ± 2.84 | 8.75 ± 3.62 |
| 105 days | 6.43 ± 4.41 | 3.57 ± 2.81 | 8.94 ± 4.04 |
| *WOMAC stiffness* |  |  |  |
| Baseline | 4.02 ± 1.82 | 3.04 ± 1.86 | 4.88 ± 1.29 |
| 5 days | 3.58 ± 1.80 | 2.54 ± 1.60 | 4.50 ± 1.44 |
| 10 days | 2.87 ± 1.85 | 1.82 ± 1.47 | 3.78 ± 1.68 |
| 15 days | 2.88 ± 1.92 | 1.68 ± 1.16 | 3.94 ± 1.85 |
| 45 days | 2.90 ± 1.95 | 1.79 ± 1.23 | 3.88 ± 1.95 |
| 75 days | 2.85 ± 1.74 | 1.89 ± 1.62 | 3.69 ± 1.40 |
| 105 days | 2.92 ± 1.95 | 1.50 ± 1.45 | 4.16 ± 1.42 |
| *WOMAC physical function* |  |  |  |
| Baseline | 29.28 ± 12.79 | 19.54 ± 9.52 | 37.81 ± 8.45 |
| 5 days | 25.95 ± 12.63 | 16.57 ± 7.33 | 34.16 ± 10.38 |
| 10 days | 21.40 ± 11.96 | 13.07 ± 7.41 | 28.69 ± 10.35 |
| 15 days | 21.97 ± 15.62 | 10.57 ± 7.91 | 31.94 ± 13.78 |
| 45 days | 23.20 ± 15.40 | 12.54 ± 8.98 | 32.53 ± 13.74 |
| 75 days | 23.85 ± 13.54 | 14.14 ± 8.81 | 32.34 ± 11.05 |
| 105 days | 23.73 ± 15.35 | 13.54 ± 9.72 | 32.66 ± 13.79 |

*Abbreviation.* NRS, Numeric Rating Scale; WOMAC, Western Ontario and McMaster Universities Osteoarthritis Index; tDCS, transcranial direct current stimulation

### **Supplemental Table 4.** Descriptive statistics for all measures throughout the study: sham tDCS

|  |  | mean ± standard deviation | |
| --- | --- | --- | --- |
|  | All | Group 1 | Group 2 |
| *NRS pain* |  |  |  |
| Baseline | 50.63 ± 21.77 | 40.09 ± 18.89 | 63.52 ± 17.96 |
| 5 days | 44.77 ± 24.48 | 31.30 ± 19.41 | 61.22 ± 1.62 |
| 10 days | 43.20 ± 22.90 | 33.39 ± 22.38 | 55.19 ± 17.40 |
| 15 days | 49.55 ± 25.74 | 36.30 ± 23.90 | 65.74 ± 17.47 |
| 45 days | 48.12 ± 27.00 | 32.64 ± 23.10 | 67.04 ± 17.93 |
| 75 days | 48.20 ± 27.36 | 35.36 ± 23.79 | 63.89 ± 23.18 |
| 105 days | 50.20 ± 28.41 | 36.52 ± 25.39 | 66.93 ± 22.59 |
| *WOMAC pain* |  |  |  |
| Baseline | 8.92 ± 3.30 | 6.85 ± 2.82 | 11.44 ± 1.67 |
| 5 days | 7.78 ± 3.82 | 5.21 ± 2.04 | 10.93 ± 3.05 |
| 10 days | 7.33 ± 4.17 | 4.61 ± 2.47 | 10.67 ± 3.31 |
| 15 days | 6.93 ± 3.91 | 4.52 ± 2.48 | 9.89 ± 3.26 |
| 45 days | 7.07 ± 3.93 | 4.52 ± 2.68 | 10.19 ± 2.79 |
| 75 days | 6.77 ± 3.65 | 4.36 ± 2.29 | 9.70 ± 2.74 |
| 105 days | 6.62 ± 3.92 | 4.24 ± 2.75 | 9.52 ± 3.12 |
| *WOMAC stiffness* |  |  |  |
| Baseline | 4.28 ± 1.63 | 3.64 ± 1.58 | 5.07 ± 1.33 |
| 5 days | 3.77 ± 1.70 | 3.12 ± 1.65 | 4.56 ± 1.42 |
| 10 days | 3.42 ± 1.44 | 2.55 ± 1.09 | 4.48 ± 1.05 |
| 15 days | 3.40 ±1.62 | 2.48 ± 1.30 | 4.52 ± 1.22 |
| 45 days | 3.40 ± 1.60 | 2.61 ± 1.34 | 4.37 ± 1.33 |
| 75 days | 3.33 ± 1.56 | 2.64 ± 1.19 | 4.19 ± 1.55 |
| 105 days | 3.10 ± 1.77 | 2.48 ± 1.48 | 3.85 ± 1.83 |
| *WOMAC physical function* |  |  |  |
| Baseline | 30.68 ± 11.63 | 22.94 ± 8.55 | 40.15 ± 6.91 |
| 5 days | 27.48 ± 12.56 | 19.18 ± 8.50 | 37.63 ± 8.65 |
| 10 days | 25.43 ± 13.09 | 16.21 ± 7.79 | 36.70 ± 8.61 |
| 15 days | 25.47 ± 12.68 | 16.61 ± 7.99 | 36.30 ± 8.04 |
| 45 days | 25.63 ± 12.92 | 17.42 ± 9.55 | 35.67 ± 8.76 |
| 75 days | 25.07 ± 11.91 | 17.36 ± 8.26 | 34.48 ± 8.40 |
| 105 days | 25.50 ± 14.10 | 17.18 ± 9.33 | 35.67 ± 12.21 |

*Abbreviation.* NRS, Numeric Rating Scale; WOMAC, Western Ontario and McMaster Universities Osteoarthritis Index; tDCS, transcranial direct current stimulation
